# COVID-19 severe pneumonia in Mexico City – First experience in a Mexican hospital

**DOI:** 10.1101/2020.04.26.20080796

**Authors:** B Valente-Acosta, I Hoyo-Ulloa, L Espinosa-Aguilar, R Mendoza-Aguilar, J Garcia-Guerrero, D Ontañon-Zurita, B Gomez-Gomez, O Fueyo-Rodríguez, JM Vera-Zertuche, RJ Anzola-Arias, JV Jiménez-Ceja, D Horta-Carpinteyro, C Olvera-Guzman, J Aguirre-Sanchez, J Franco-Granillo, L Jauregui-Camargo, E Sada-Díaz, R Saavedra-Perez-Salas, A Palomar-Lever, F Moreno-Sánchez

## Abstract

**Background:** Coronavirus Disease 2019 (Covid-19) pandemic since its first confirmed case, has changed the world. The need for accurate and truthful information is vital. Mexico and Latin America have been widely affected, so having local epidemiological data, will be of great clinical utility.

**Methods:** A total of 33 hospitalized patients with Covid-19 pneumonia (either severe or critical) were identified from electronic health record in a third level care private hospital in Mexico City from March 13rd to April 13rd, 2020. We conducted a descriptive study of patients for characterization of the clinical, laboratory and radiologic findings, as well as complications.

**Results:** The mean age was 60.6±12.68 years and 23 (69.7%) were males. Twenty-three patients (69.6%) were overweight or obese. The median duration of symptoms before admission was 7 days. All the patients required mechanical invasive ventilation. The median duration of the mechanical ventilation was 12±2.6 days and all patients were extubated except one. All patients were started on antiviral treatment in the first 24 hours after admission once the diagnosis of Covid19 pneumonia was made. There was no difference between the treatment option and the length of stay. The extubation rate was higher (91.6%) than in other series, with no fatalities even though they were treated with different regimens.

**Conclusions:** This one-centre experience describes the epidemiology, treatment and outcome of 33 patients with severe or critical COVID pneumonia admitted to the ICU. Most patients in our series were overweight or obese male, which we observed were of higher risk to present critical pneumonia, as well as high levels of Interleukin-6. The foregoing is relevant, due to the high incidence of these comorbidities in our country.

## Background

It is well known that since the first cases were identified in December 2019 in Wuhan, the capital of China’s Hubei province, coronavirus disease 2019 (COVID-19) has changed not only medical practice but also the world itself. The virus that causes this disease has been named severe acute respiratory syndrome coronavirus 2 (SARS-CoV-2) because of its resemblance to SARS-CoV: both belong to the coronavirus family.(1)

As the pandemic evolves, having accurate local and international information has become vitally important. To the best of our knowledge, there are no published data on the clinical presentation and epidemiology of this disease in the Mexican population. The flow of information from China, Europe and the United States could be used as a basis for our population; however, we must recognise that the Mexican population has very different social and health characteristics.(2)

We present a retrospective case series from a tertiary-level private medical centre in Mexico City, which we believe is of great value, since it represents one of the first experiences in Latin America of the clinical behaviour of the virus.

## Methods

### Setting and study population

We report a case series of patients with severe and critical COVID-19 pneumonia admitted to the American British Cowdray (ABC) Medical Center from 13 March to 13 April 2020. The ABC hospital is a tertiary-level care private hospital in Mexico City.

Thirty-three adults (≥ 18 years) were identified and divided into two groups (severe or critical pneumonia) depending on the pneumonia severity according to the Chinese Clinical Guidance for COVID-19 Pneumonia Diagnosis and Treatment.(3) Patients were classified in one of these groups depending on their presentation in the first 48 hours after admission. Pregnant women and children younger than 18 years of age were excluded from the study.

### Data sources

We obtained demographic, clinical, laboratory and radiologic data at admission and during the patients’ hospitalisation from the electronic health record. The laboratory data and information on the treatment given were collected for all patients up to the time of the data cut, which occurred on 19 April 2020. Data were anonymised before analysis. Informed consent was waived. The institutional ethics and research committee approved the protocol.

### Study definitions

A confirmed case of COVID-19 was defined by a positive result on a reverse transcriptase–polymerase chain reaction (RT-PCR) assay of a specimen collected on a nasopharyngeal swab or a patient presenting with clinical and radiological signs compatible with COVID-19 despite at least two consecutive negative Sars-CoV-2 RT-PCR determinations.

We divided patients into two groups: 1. Severe disease was defined as patients presenting with dyspnoea, a respiratory rate of more than 30/min, peripheral capillary oxygen saturation (SpO_2_) of 90% breathing ambient air and/or a PaO_2_/Fio_2_ ratio less than 231 (in agreement with the Mexico City altitude); 2. Critical disease was defined as severe pneumonia with respiratory failure requiring invasive mechanical ventilation, shock and/or other organ dysfunctions requiring admission to the intensive care unit (ICU).

### Specimen collection and testing

Clinical specimens for SARS-CoV-2 diagnostic testing were obtained in accordance with the Center for Disease Control and Prevention guidelines. Different kits targeted the SARS-CoV-2 E-gene and the RdRP gene, including the RNeasy Mini Kit (Qiagen), and the LightCycler II Z480 (Roche®) with LightMix Modular detection system (TIB Molbiol, Roche) targeted the CoV E-gene, the CoV N-gene and the CoV RdRP gene.(4) Treatment was chosen at the attending physician’s discretion. All the patients or patients’ families gave their informed consent for the compassionate use of these drugs. Afterwards, and only for analysis purposes, the patients were classified according to the medications received into the following groups:

1. Lopinavir/ritonavir (LPV/r) 400 mg/100 mg twice daily (BID) for 7 days + Interferon beta-1b (IFNb-1b) 0.25 mg every 48 h for 3 to 7 doses + Azithromycin (AZI) 500 mg initial dose and 250 mg daily for 5 days.
2. LPV/r 400 mg/100 mg (BID) for 7 days + IFNb-1b 0.25 mg every 48 h for 3 to 7 doses + AZI 500mg initial dose and then 250 mg daily for 5 days + Hydroxychloroquine (HCQ) loading dose of 400 mg BID and then 200 mg BID for 5 to 10 days.
3. LPV/r 400 mg/100 mg BID for 7 days + AZI 500 mg initial dose and then 250 mg daily for 5 days + HCQ loading dose of 400 mg BID and then 200 mg BID for 5 to 10 days.
4. AZI 500 mg initial dose and then 250 mg daily for 5 days + HCQ loading dose of 400 mg BID and then 200 mg 3 times a day (TID) for 5 to 10 days.

Tocilizumab (400 mg IV) could be added to one of the previous regimes if the patient developed clinical and radiological signs of deterioration despite current treatment. Patients had to have a high interleukin-6 (IL-6) level. Before the tocilizumab infusion, patients had a complete assessment, including an electrocardiogram, HIV test, viral hepatitis panel and QuantiFERON-TB Gold.

### Statistical analysis

We used descriptive statistics expressed as numbers (percentages) for the categorical variables, and the continuous variables were expressed as mean ± standard deviation (SD) and median with interquartile range (IQR) values in accordance with their distribution. The Student’s t-test or the Mann-Whitney test were performed to compare the differences between the continuous variables according to their distribution. The Kolmogorov–Smirnov test was used as evidence of normality. Significant differences between the categorical variables were evaluated using the Chi-square test. The ANOVA test was used to evaluate the differences between different treatment schemes and the length of stay. A p-value less than 0.05 was considered statistically significant. All statistical analyses were performed using SPSS version 22.0 (IBM Corp., Armonk, NY, USA).

## Results

### Patient characteristics

We included all the patients managed by our infectious diseases team that were admitted to the intensive care COVID ward in the month following the first admission, which was on 12 March 2020. All the patients had severe or critical pneumonia. Patients were in the care of the intensive care and infectious diseases teams. The clinical characteristics of the patients at admission are depicted in Table 1. We had 25 patients with severe pneumonia at admission and 8 patients with critical pneumonia that developed in the first 48 hours after admission.

**Table 1.** Clinical characteristics of the patients at admission and treatment given.

|  | **Severe**<br>**Pneumonia**<br>**n = 25** | **Critical**<br>**Pneumonia**<br>**n = 8** | **p** |
| --- | --- | --- | --- |
| Mean age | 59.38 ± 13.23 | 64.63 ± 9.25 | NS |
| **Sex – n (%)** |  |  |  |
| Male | 17 (68) | 6 (75) | NS |
| Female | 8 (32) | 2 (25) |  |
| Mean body mass index | 26.91 ± 4.11 | 30.16 ± 6.21 | NS |
| SARS-CoV2 positive RT-PCR<br>n (%) | 22 (88) | 7 (87.5) | NS |
| **Symptoms – n (%)** |  |  |  |
| Fever | 20 (80) | 5 (62.5) | NS |
| Cough | 16 (64) | 5 (62.5) | NS |
| Diarrhoea | 6 (24) | 2 (25) | NS |
| Headache | 10 (40) | 4 (50) | NS |
| Malaise | 22 (88) | 6 (75) | NS |
| Median respiratory rate (IQR) | 22 (20–23) | 25.5 (22–29.5) | < 0.05 |
| Mean PaO<sub>2</sub>/FiO<sub>2</sub> ratio | 217 ± 31.2 | 122 ± 66.1 | < 0.05 |
| **Comorbidities – n (%)** |  |  |  |
| Any | 16 (64) | 7 (87.5) | NS |
| Overweight or Obesity | 15 (60) | 7 (87.5) | NS |
| Smoking | 10 (38.5) | 3 (37.5) | NS |
| Hypertension | 7 (28) | 5 (62.5) | NS |
| ACE or ARB treatment | 5 (20) | 4 (50) | NS |
| Diabetes mellitus | 6 (24) | 2 (25) | NS |
| Cardiopathy | 3 (12) | 0 (0) | NS |
| COPD | 3 (12) | 1 (12.5) | NS |
| Immunosuppression | 1 (4) | 1 (12.5) | NS |
| Median duration of symptoms<br>before admission – days (IQR) | 7 (4–10) | 6.5 (3.5–9.5) | NS |
| **Treatment\* – n (%)** |  |  |  |
| LPV/r + IFNb-1b + AZI | 2 (8) | 2 (25) | NS |
| LPV/r + IFNb-1b +HCQ + AZI | 6 (24) | 3 (37.5) | NS |
| LPV/r + HCQ + AZI | 6 (24) | 3 (37.5) | NS |
| HCQ + AZI | 11 (44) | 0 (0) | < 0.05 |
| Tocilizumab in addition to previous treatment | 11 (44) | 6 (75) | NS |

**IQR: interquartile range, NS: not statistically significant, PaO<sub>2</sub>/FiO<sub>2</sub>∴, RT-PCR: reverse transcriptase–polymerase chain reaction.**
**\*LPV/r: Lopinavir/ritonavir, IFNb 1b: Interferon beta-1b, HCQ: Hydroxychloroquine, AZi: Azithromycin.**

The patients’ mean age was 60.6 ± 12.68 years, and 23 (69.7%) were males. The median duration of symptoms before admission was 7 (IQR 5–8) days. Most (90.9%) of the patients had at least one nasopharyngeal positive RT-PCR for SARS-CoV2. Twenty-eight (84.8%) patients had malaise, 25 (75.8%) had fever, 21 (63.6%) had a cough, 14 (42.4%) had headache, and only 8 (24.2%) had diarrhoea at admission. One patient presented with rhabdomyolysis that was managed with aggressive fluid resuscitation. Of the 33 patients, 31 had a PCR for influenza A/B or a multiplex PCR panel for respiratory pathogens. Two patients were co-infected with influenza and one with rhinovirus.

Patients in the severe pneumonia group (SPG) had a lower respiratory rate and a higher PaO_2_/FiO_2_ ratio compared to patients in the critical pneumonia group (CPG) (22 vs 25.5, p < 0.05 and 217 ± 31.2 vs 122 ± 66.1 p, < 0.05, respectively). Twenty-three (69.7%) patients had any comorbidity. However, we did not find any significant difference between chronic medical conditions among both groups. Twenty-three (69.6%) patients were overweight or obese. Thirteen (39.3%) patients were current smokers. Twelve (35.3%) patients had hypertension, and only 9 were being treated with ACE or ARB drugs.

All patients were started on antiviral treatment in the first 24 hours after admission once the diagnosis of COVID-19 pneumonia was made. Four patients were treated with lopinavir/ritonavir and interferon beta-1b, and 9 patients were treated with this combination plus hydroxychloroquine. Nine patients were treated with lopinavir/r and hydroxychloroquine, and eleven patients were treated with only hydroxychloroquine. All the patients were on azithromycin. There was no difference between the treatment options and the length of stay. Tocilizumab was given to 17 (51.5%) patients: 6 (75%) patients in the CPG and 11 (44%) in the SPG, including 4 patients who progressed to critical pneumonia after 48 hours of admission. On average, patients received tocilizumab 3.3 ± 2.2 days after admission.

### Laboratory and radiologic findings

Table 2 shows the laboratory and radiologic findings in the patients on admission to the hospital. All the patients had a complete blood count, a complete biochemistry panel and tests for D-dimer, ferritin, troponin I, C-reactive protein, procalcitonin and IL-6 levels. Ferritin, lactic dehydrogenase and IL-6 levels were significantly more elevated in critical pneumonia patients than in the severe pneumonia patients.

**Table 2.** Laboratory data at hospital admission and radiology findings.

|  | **Severe Pneumonia**<br>**Patients n = 25** | **Critical**<br>**Pneumonia**<br>**Patients n = 8** | **p** |
| --- | --- | --- | --- |
| White blood cell per mm<sup>3</sup><br>Median (IQR) | 4.8 (4.15–8.1) | 8.35 (4.87–14.65) | NS |
| Lymphocyte count per mm<sup>3</sup><br>Mean (SD) | 1.01 (0.49) | 1.21 (0.69) | NS |
| Creatinine mg/dL<br>Median (IQR) | 0.99 (0.75–1.14) | 0.91 (0.78–1.68) | NS |
| Ferritin mg/dL<br>Median (IQR) | 776 (379–1413) | 2104 (1065.5–2937) | < 0.05 |
| Lactic dehydrogenase mg/dL<br>Mean (SD) | 262.27 (92.64) | 381.57 (125.95) | < 0.05 |
| D-Dimer mg/dL<br>Mean (SD) | 796.8 (447.99) | 1022.62 (359.01) | NS |
| Troponin I mg/dL<br>Median (IQR) | 11.9 (7.55–16.1) | 29.25 (10.95–37.5) | NS |
| IL-6 mg/dL<br>Median (IQR) | 69.85 (15.97–121.32) | 199.5 (77.75–231.25) | < 0.05 |
| C-reactive protein mg/dL<br>Mean (SD) | 9.31 (8.14) | 13.15 (10.39) | NS |
| Procalcitonin mg/dL<br>Mean (SD) | 1.01 (3.9) | 0.80 (1.13) | NS |
| **CT findings – n (%)** |  |  |  |
| Bilateral ground-glass opacification | 12 (48) | 0 (0) | < 0.05 |
| Bilateral alveolar infiltrates | 1 (4) | 2 (25) |  |
| Bilateral mixed infiltrates (ground-glass and alveolar) | 12 (48) | 6 (75) |  |

CT: computed tomography, IL6: interleukin-6, IQR: interquartile range, NS: not statistically significant, SD: standard deviation.

All the patients had bilateral infiltrates in the computed tomography scan (CT-scan). Twelve (48%) patients with severe pneumonia had bilateral ground-glass infiltrates, while 6 (75%) patients with critical pneumonia had bilateral mixed infiltrates (alveolar occupation and ground-glass infiltrates).

### Critical patients

Eight patients had critical pneumonia or developed it in the first 48 hours after admission. Four (16%) patients in the SPG developed critical pneumonia 48 hours after admission. All the critical patients required mechanical invasive ventilation. It is worth mentioning that we adhere to the Berlin definitions regarding the severity of respiratory failure, and if clinical situation required it, alternatives modalities of mechanical ventilation were implemented, including the prone position. The median duration of the mechanical ventilation was 12 ± 2.6 days. All the patients were extubated except one. The patient who could not be extubated had previous Chronic obstructive pulmonary disease (COPD) and is now awake with a tracheostomy. Among the 12 patients who had critical pneumonia, 10 (83.3%) received tocilizumab.

### Complications

We only documented two drug-related adverse reactions. One patient stopped lopinavir/ritonavir on the fifth day because of a considerable increase in bilirubin. After discontinuation, the bilirubin came down in 3 days. Another patient stopped azithromycin because of atrial fibrillation and a prolonged QT interval.

All the patients who required invasive mechanical ventilation had at least one bronchoalveolar lavage (BAL) for bacterial and fungal cultures. Three patients developed ventilator-associated pneumonia with extended spectrum beta-lactamase *Escherichia coli, Enterococcus faecalis* and *Stenotrophomonas maltophilia*. One patient developed invasive pulmonary aspergillosis, diagnosed by a positive galactomannan in BAL, although we only performed a galactomannan testing of BAL fluid in 4 patients who had invasive mechanical ventilation.

Two patients developed subsegmental pulmonary emboli despite low-molecular-weight heparin thromboprophylaxis. Two patients required re-intubation, one because of life-threatening abdominal bleeding that was treated by vascular surgery and another who developed complete right lung atelectasis. Both patients were able to be extubated in the next 72 hours after the bleeding and atelectasis resolved.

### Outcomes

Table 3 depicts the major outcomes of our cases. All patients were followed up until hospital discharge. The median follow-up day was 8 (IQR 5.5–19). For the 9 (27%) patients still in the hospital, we have at least 6 days of follow-up. As of 18 April 2020, none of the 33 patients has died. Twenty (80%) patients in the SPG and 4 (50%) patients in the CPG have been discharged. The median length of stay was 7.5 days (4.7–8.7) for the SPG and 23 days (16–24.7) for the CPG.

**Table 3.**
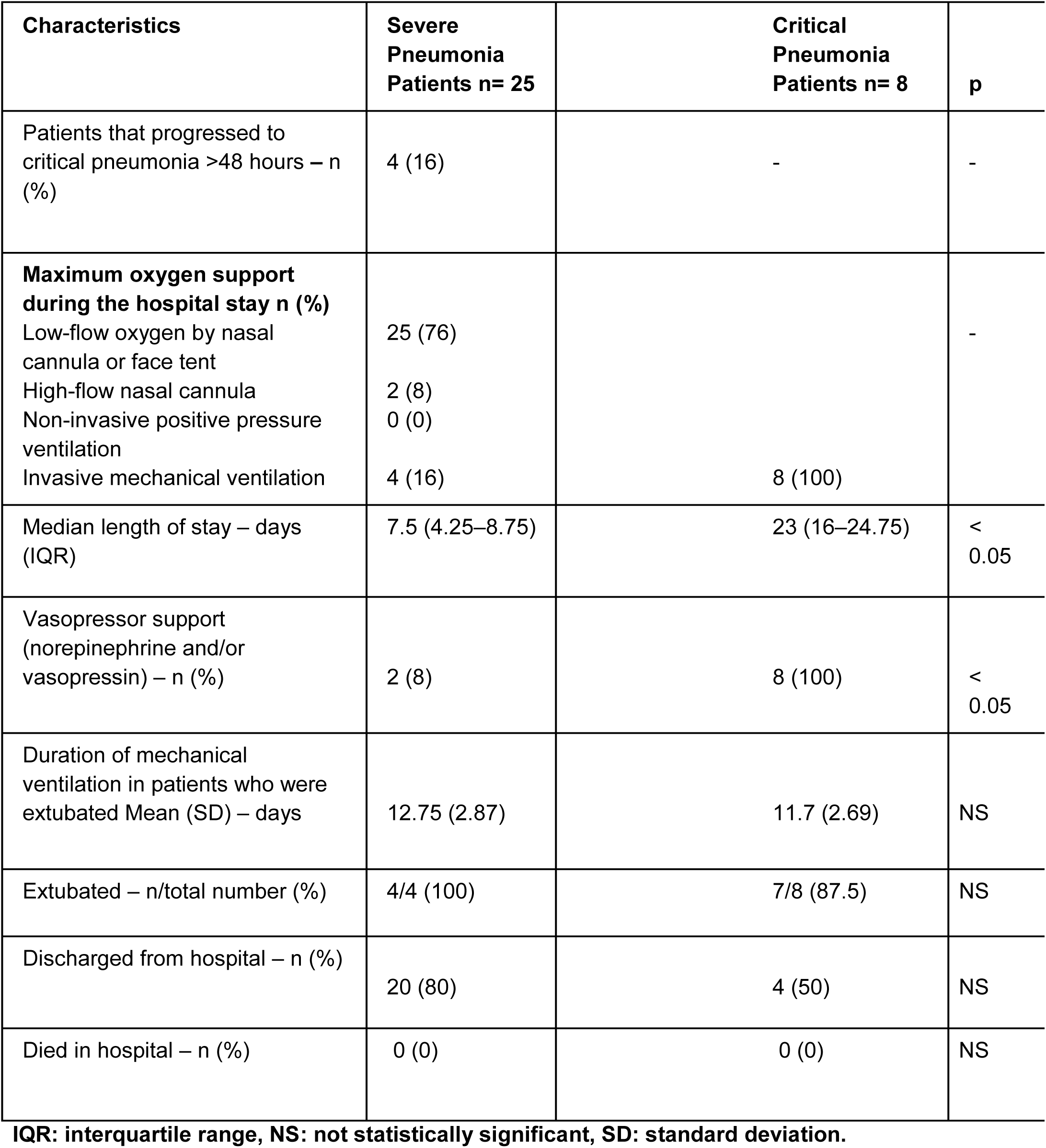
Outcomes.

| Characteristics | Severe<br>Pneumonia<br>Patients n= 25 | Critical<br>Pneumonia<br>Patients n= 8 | p |
| --- | --- | --- | --- |
| Patients that progressed to critical pneumonia >48 hours – n (%) | 4 (16) | - | - |
| **Maximum oxygen support during the hospital stay n (%)** |  |  | - |
| Low-flow oxygen by nasal cannula or face tent | 25 (76) |  |  |
| High-flow nasal cannula | 2 (8) |  |  |
| Non-invasive positive pressure ventilation | 0 (0) |  |  |
| Invasive mechanical ventilation | 4 (16) | 8 (100) |  |
| Median length of stay – days (IQR) | 7.5 (4.25–8.75) | 23 (16–24.75) | < 0.05 |
| Vasopressor support (norepinephrine and/or vasopressin) – n (%) | 2 (8) | 8 (100) | < 0.05 |
| Duration of mechanical ventilation in patients who were extubated Mean (SD) – days | 12.75 (2.87) | 11.7 (2.69) | NS |
| Extubated – n/total number (%) | 4/4 (100) | 7/8 (87.5) | NS |
| Discharged from hospital – n (%) | 20 (80) | 4 (50) | NS |
| Died in hospital – n (%) | 0 (0) | 0 (0) | NS |

IQR: interquartile range, NS: not statistically significant, SD: standard deviation.

## Discussion

This single-centre experience describes the epidemiology, treatment and outcome of 33 patients with severe or critical COVID-19 pneumonia admitted to the ICU between 12 March and 13 April 2020.

Most patients in our case series were overweight or obese males with a chronic illness, most commonly hypertension or diabetes, similar to other reports.(1,5) Obesity and metabolic syndrome are chronic inflammatory diseases that make patients more prone to infectious complications and are known to have greater mortality in COVID-19 patients than patients with normal body mass index (BMI).(6) In our case series, all but one of the patients in the critical group were overweight or obese, and there was a trend towards higher BMI in the critical group compared to those in the severe group. The mean BMI in both groups was higher than those reported previously by Liu et al.(7) This finding is expected, since the prevalence of overweight and obesity in Mexico is one of the highest in the world, with 75.2% of the adult population living with a BMI above 25.(8) The pathophysiologic basis behind the more severe clinical picture of COVID-19 in patients with obesity seems to be a chronic inflammatory and prothrombotic state, higher ACE2 concentrations in the alveolar epithelium plus a compromised pulmonary physiology.(9) Moreover, adipose tissue has been known to be a reservoir for some viruses, such as HIV and CMV; its role as a tissue reservoir in SARS-CoV-2 remains to be studied.(10)

Fever and cough were the most common presenting symptoms, with a mean duration of 7 days before hospital admission. A higher respiratory rate and a lower PaO_2_/ FiO_2_ ratio at admission were significantly associated with the development of critical pneumonia. These are two well-known markers of serious respiratory illness.(11)

We found only two patients with influenza and one with a rhinovirus coinfection. Most of our patients were tested for other respiratory pathogens by molecular analysis. In contrast with other series that describe up to 20% of respiratory virus coinfection.(12)

As described in Chinese and Italian reports, higher ferritin and lactic dehydrogenase levels at admission were also significantly associated with critical pneumonia.(13,14) Moreover, we found that higher IL-6 concentration was significantly associated with critical pneumonia and the need for mechanical ventilation. This finding has previously been reported.(15,16)

All our patients were treated with different regimens of potential antiviral drugs against SARS-CoV2. The medication was started within 24 hours of admission. We could not compare the different schemes because our report is only a case series compared to well-designed trials.(17,18)

Tocilizumab was given to 11 (44%) and 6 (75%) of our severe and critical pneumonia patients, respectively. The rationale for such a decision was based on observational data implicating the overwhelming systemic inflammatory cascade as a culprit in the physiopathology of this new and poorly understood disease and the plausible role for immunomodulation in these populations.(19,20)

All the patients with critical COVID-19 pneumonia required mechanical ventilation with a mean duration of 12 days, similar to critically ill patients in Seattle, where the duration of mechanical ventilation was 10 days.(21) Noteworthy is that our extubation rate was higher (91.6%) than in other series. In addition, we did not have any fatalities.

Regarding complications, there were 3 cases of ventilator-associated pneumonia and only one with a multi-drug resistant bacterium, an extended spectrum beta-lactamase *E. coli*. Secondary bacterial infection has been related to longer hospital stays and worse outcomes.(13) Only one patient was diagnosed with probable invasive pulmonary aspergillosis, a fungal coinfection described in another series.(22)

Pulmonary embolism was observed in two patients, despite receiving thromboprophylaxis. Severe COVID-19 pneumonia can be complicated with prothrombotic coagulopathy, causing both major thromboembolic events and microthrombi in end-organ capillary beds. Therefore, it is currently recommended that all patients (unless contraindicated) should receive thromboprophylaxis, and those with elevated coagulation markers (specifically D-dimer) should receive full dose anticoagulation, as it appears to be associated with lower mortality. Our patients were managed following these recommendations.(23,24)

Our study has several limitations: it is a small descriptive case series report, and there may be confounders in the analysis of the results, in concordance with most described data in the current literature. Another disadvantage is the lack of generalisability and the fact that 9 patients remained in the hospital at the time of data censoring on April 19, 2020.

## Data Availability

The data that support the findings of this study are available from the corresponding author, upon reasonable request.

